# Phenoflow: A Microservice Architecture for Portable Workflow-based Phenotype Definitions

**DOI:** 10.1101/2020.07.01.20144196

**Authors:** Martin Chapman, Luke V. Rasmussen, Jennifer A. Pacheco, Vasa Curcin

## Abstract

Phenotyping is an effective way to identify cohorts of patients with particular characteristics within a population. In order to enhance the portability of a phenotype definition across institutions, it is often defined abstractly, with implementers expected to realise the phenotype computationally before executing it against a dataset. However, unclear definitions, with little information about how best to implement the definition in practice, hinder this process. To address this issue, we propose a new multi-layer, workflow-based model for defining phenotypes, and a novel authoring architecture, Phenoflow, that supports the development of these structured definitions and their realisation as computable phenotypes. To evaluate our model, we determine its impact on the portability of both code-based (COVID-19) and logic-based (diabetes) definitions, in the context of key datasets, including 26,406 patients at North-western University. Our approach is shown to ensure the portability of phenotype definitions and thus contributes to the transparency of resulting studies.

## Introduction

Learning Health Systems require high-quality, routinely collected electronic health record (EHR) data to drive analytics and research, and translate the outputs of novel techniques such as machine learning into patient care and service improvement. To achieve this, the data used for research need not only be of high-quality, but methods associated with its use need be transparent and reproducible to ensure that any findings can be validated by the research community and generalised to other populations. At the core of this challenge is the ability to reliably identify clinically equivalent research-grade patient cohorts. These cohorts are precise enough to conduct meaningful research by identifying individuals with a particular disease, sets of comorbidities, medical histories, a demographic profile or any other relevant patient-specific information – a process known as EHR-based phenotyping^1^.

The popularity of EHR data for research has increased the documentation and sharing of phenotypes derived from research datasets in order to stimulate reuse, reduce variation in phenotype definitions across data sources, and ultimately simplify and support the identification of clinically equivalent populations for research and healthcare applications. The reuse of existing phenotype definitions necessitates the ability to discover and access curated and validated phenotype definitions. Pioneering efforts in building standardised phenotype repositories, such as the Phenotype Knowledge Base (PheKB), CALIBER, Million Veterans Program (MVP) and *All of Us* consortium have achieved notable success within their research programmes, with thousands of registered usages^2,3^.

In an attempt to ensure the portability of a phenotype across multiple research use cases, the logic that comprises a phenotype definition is often represented abstractly within these repositories, where it is structured as, for example, a list of codes (e.g. Figure 1), or as a data flow diagram (e.g. Figure 2). This abstract representation is designed to guide the development of a computable form of the phenotype, such as an executable script or a data pipeline, for a particular use case. However, in practice, the portability of these definitions is often low: a lack of clarity in the abstract definition, either in terms of terminology or structure, make them hard to interpret in order to produce a computable form, and the technical skill burden on the computable phenotype author is high, as the abstract nature of each definition means that little is communicated about the realisation of each phenotype in practice.

**Figure 1:**
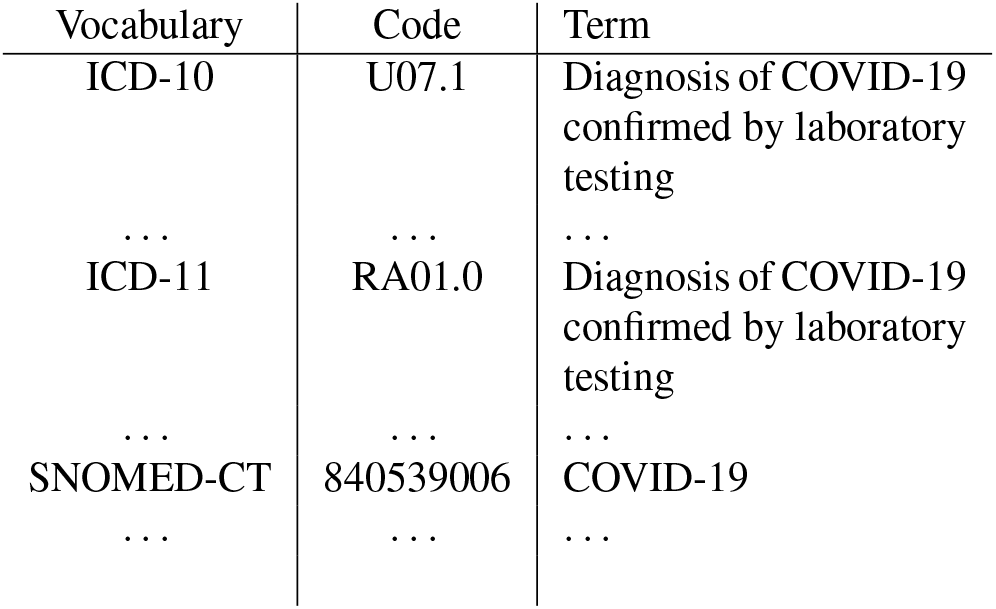
Extract of code lists used for defining COVID-19 patients in an EHR system (Source: http://covid19-phenomics.org/).

**Figure 2:**
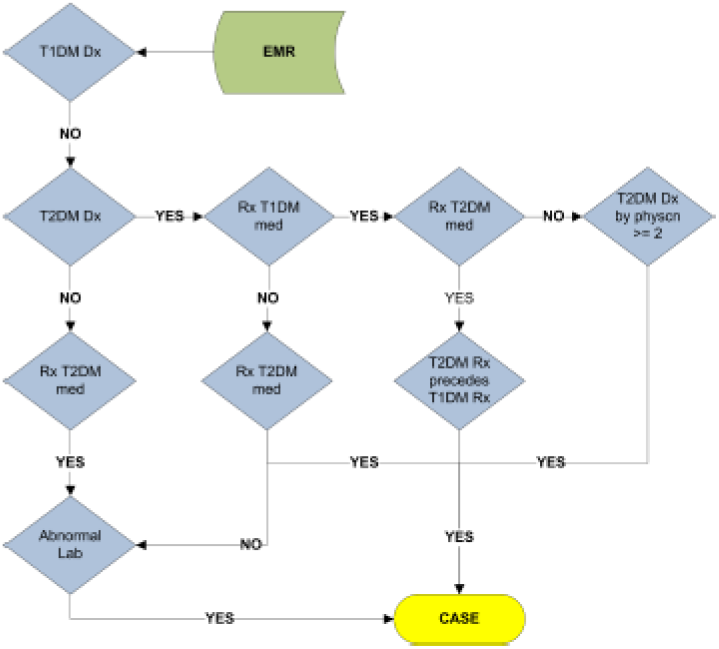
Data flow for defining T2DM patients in an EHR system (Source: https://phekb.org/phenotype/type-.2-diabetes-mellitus).

## Methods

In order to address the difficulty of deriving computable forms from phenotype definitions, we propose a novel phenotype definition model, which aims to increase portability by improving clarity, and more explicitly defining the structure of computable forms. The formulation of the proposed model was based on the experiences of initiatives such as the UK eScience and US Cyberinfrastructure programmes^4^, which developed *scientific workflow* models for orchestrating and coordinating their computational tasks. In addition, the functional (re-)modelling of different phenomena in a number of different domains was used as a basis for the proposed model – in particular, work in *hierarchical modelling*; the representation of a phenomena at different levels of abstraction^5^, e.g. in bioinformatics software architectures^6^. Finally, the authors themselves have developed a number of different models as part of prior studies, including work on complex systems^7^, semantics for hierarchical composition^8^ and phenotyping from large scale EHR repositories^9^.

In order to evaluate how our new *structured definition* model impacts portability, we first collected a set of 278 existing phenotype definitions from a number of different phenotype repositories, including PheKB (https://phekb.org) and CALIBER (e.g. https://portal.caliberresearch.org). We then re-authored these definitions according to our model, and used them to produce corresponding computable forms. In examining these definitions, we identified that they fall broadly into two categories: *code-based* definitions that identify patient cohorts using a list of clinical codes, and *logic-based* definitions that identify patients using a series of logical statements. To evaluate the impact of our model on the portability of the phenotype definitions in each of these categories, we selected a representative phenotype from each category, including a code-based Coronavirus disease 2019 (COVID-19) definition (Figure 1) and a logic-based Type 2 Diabetes Mellitus (T2DM) definition (Figure 2), and compared the portability of each re-authored definition with the original, using the *Knowledge conversion, clause Interpretation, and Programming* (KIP) phenotype portability scoring system^10^.

Prior to applying the KIP scoring system, it was important to verify that our re-authoring approach resulted in structured definitions that still captured the required phenotype logic. To do this we executed the computable forms derived from the original definitions of our representative phenotypes and the computable forms derived from the structured definitions of these phenotypes against a given patient dataset, and verified that the same patients were identified by both implementations. For example, as a part of our evaluation, the phenotype definition for COVID-19 was obtained from CALIBER (http://covid19-phenomics.org), re-authored by one of the authors (MC), and a computable form produced from both the original definition (as one did not exist) and the structured definition. Similarly, the definition for T2DM, and its corresponding Konstanz Information Miner (KNIME) pipeline implementation (Figure 3), were obtained from PheKB (https://phekb.org/phenotype/type-2-diabetes-mellitus). This definition, like the definition for COVID-19, was then re-authored by one of the authors (MC), and a new computable form was produced. The original COVID-19 implementation and the new computable form were then executed against a cohort of 1468 individuals who tested positive for COVID-19 at Guy’s and St. Thomas’ NHS Foundation Trust (GSTT), London, while the T2DM implementations were executed against a cohort of 26,406 possible T2DM patients, taken from the Northwestern Medicine Enterprise Data Warehouse (NMEDW), as well as against publicly available data from PheKB. In the case of T2DM, the execution of the computable form derived from the structured definition against these datasets is possible because it uses the same data input format as the original KNIME pipeline implementation, and similarly creates an output file in the same structure. The GSTT dataset included a subset (*n* = 1153 cases) of hospitalised COVID-19 patients, while the NMEDW dataset included a subset (*n* = 23 cases) of patients with T2DM that had previously undergone manual chart review, both of which acted as the gold standard to validate our algorithm against. In both cases, the results of executing the structured implementation were compared with the results of executing the original implementations to confirm the same exact cases and controls were found across their respective datasets.

**Figure 3:**
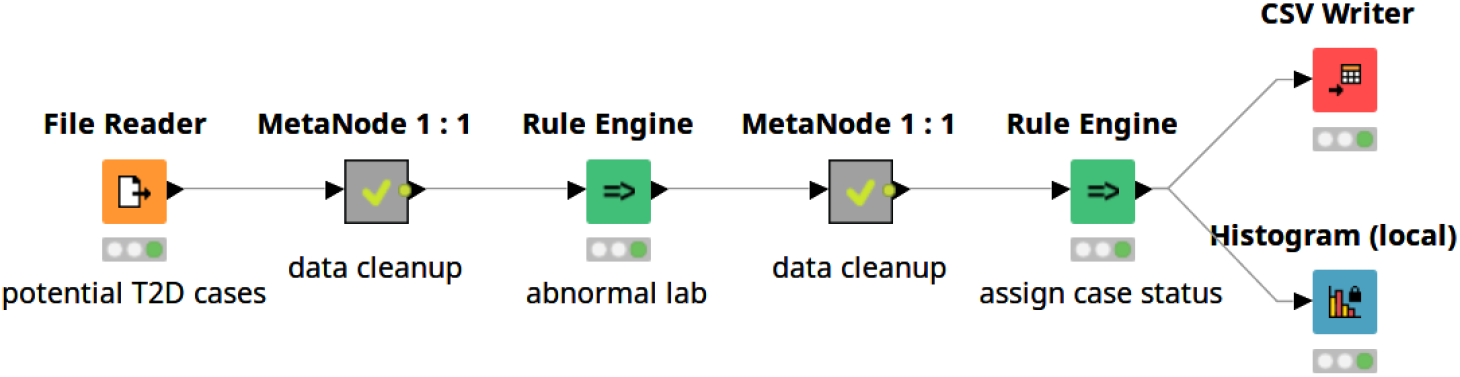
Original T2DM phenotype, implemented as the nodes of a KNIME pipeline.

### Structured Phenotype Definition

The structured phenotype definition model developed consists of a set of *layers*: abstract, functional and computational. A graphical overview of our model is given in Figure 4. Like traditional definitions, the **abstract layer** of a structured phenotype definition holds the logic of the phenotype. However, the abstract layer in our model is defined by two distinct features. Firstly, like the workflow models upon which our model is based, this layer consists of a number of sequential steps, each of which defines a single operation against a target dataset. However, steps may also be grouped, allowing for their functionality to be summarised by a single parent step. The second feature of this layer is a multi-dimensional description of each step, which consists of an ID, designed to summarise the purpose of the step using relevant clinical terminology; a longer description of the step, designed to offer a non-technical description of the logic of the step; and a categorisation of the logic of the step as an entity within a given concept ontology (broadly based on the axioms of the Phenotype Execution and Modelling Architecture (PhEMA) authoring tool (PhAT)^11^): *load* (loading data from a datasource), *logic* (generic logic to identify patients), *boolean* (boolean logic to identify patients) and *output* (the output of patients that exhibit a given phenotype to a specified location, e.g. disk). As a part of our model, we also constrain the type of logic that a step may have, depending upon its position. For example, the first step may only be of a *load* type, while the last may only be of an *output* type.

**Figure 4:**
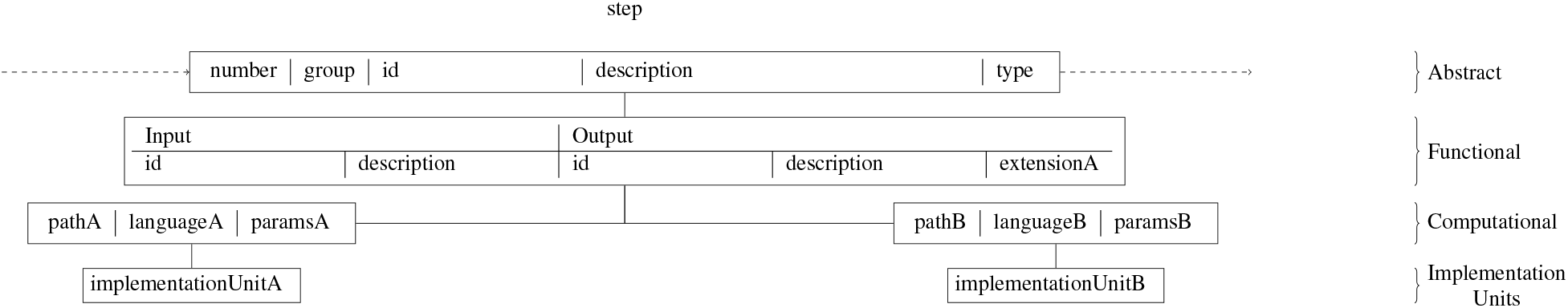
Structured phenotype definition model (step) and implementation units.

The **functional layer** of a structured phenotype definition is used to augment the information held in the abstract layer by adding metadata information about the inputs and outputs of each step. This explicit specification is an application of a functional programming paradigm^12^, and includes identifiers for both the inputs and outputs, summarising their purpose using relevant clinical terminology; a longer description of both the inputs and output to each step, designed to offer a non-technical insight; and syntactical commitments for step output (e.g. file type).

Finally, the modular **computational layer** of a structured phenotype definition is used to describe the presence of one or more implementation units (e.g. a script, data pipeline module, etc.) for each (nested) step in the abstract (and functional) layers. This description includes information about the execution environment used to run the implementation unit, and how the unit is linked to that environment.

### Generating computable phenotypes

In order to assist in the development of a computable phenotype from a structured phenotype definition, we develop a microservice architecture, *Phenoflow* (Figure 5). Software designed as a microservice architecture provides functionality based on the interactions between individual services. As a specialised type of service-oriented architecture, the microservice approach dictates that each service should only deliver one specific piece of system functionality, making it easier to achieve quality attributes such as scalability and resilience in practice^13^. This paradigm has been successfully used to structure software in several health domains, including the representation of clinical guidelines^14^.

**Figure 5:**
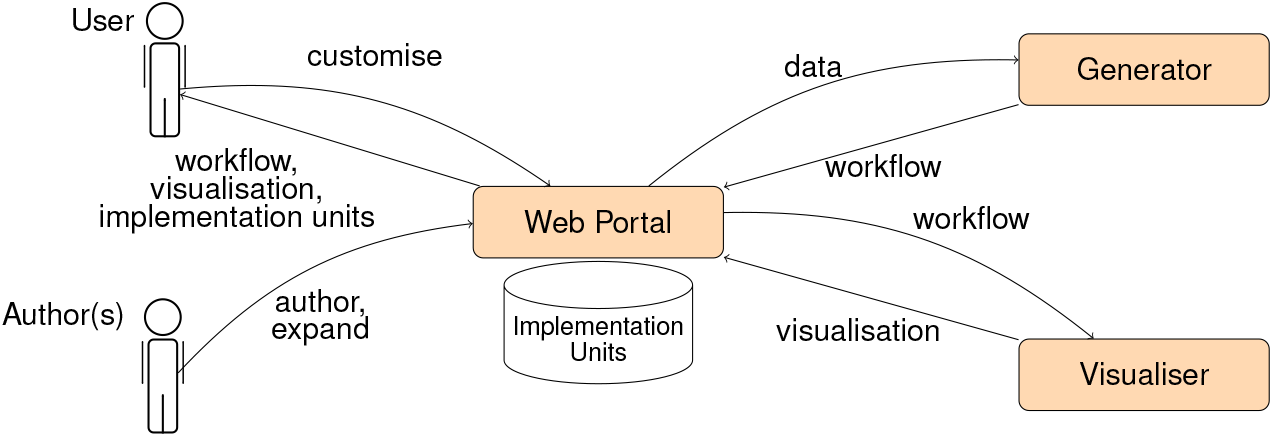
The microservices (web portal, generator and visualiser) that constitute the *Phenoflow* architecture.

Phenoflow first allows a researcher to graphically author a definition through a web portal service, where they express each step of their new definition at the abstract and functional layers by selecting the type of each step, and then by labelling, describing and, if they wish to, grouping those steps, as well as describing their inputs and outputs. This process is represented in Figure 6, where a user is in the process of defining a new boolean expression, within their abstract layer, having already defined an initial data read from an Observational Medical Outcomes Partnership (OMOP) common data model (CDM) database, another piece of logic, and the fact that a CSV file is passed between these two steps. Authors can also (indirectly) reuse the definitions produced by others in the definition of their own phenotypes. This is common in the development of new definitions that contain the same logic, but target a different data source. For example, two definitions that differ only in *load* components that separately target the OMOP CDM and an Informatics for Integrating Biology and the Bedside (i2b2) dataset.

**Figure 6:**
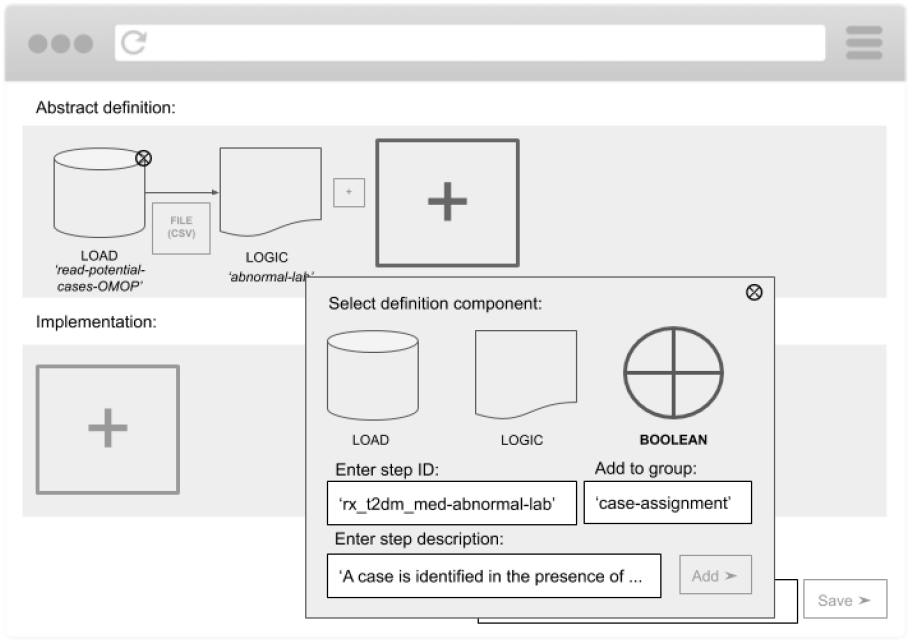
Visually defining the abstract and functional layers of a definition.

Following the specification of one or more steps in the abstract and functional layers, the researcher graphically connects each step to an implementation unit (e.g., a Python script, or a KNIME module; their choice across the steps does not have to be homogeneous), which they supply to the portal, in order to generate the computational layer. This process is represented in Figure 7, where the author is uploading the module of KNIME pipeline as the implementation counterpart of their priorly defined boolean expression step.

**Figure 7:**
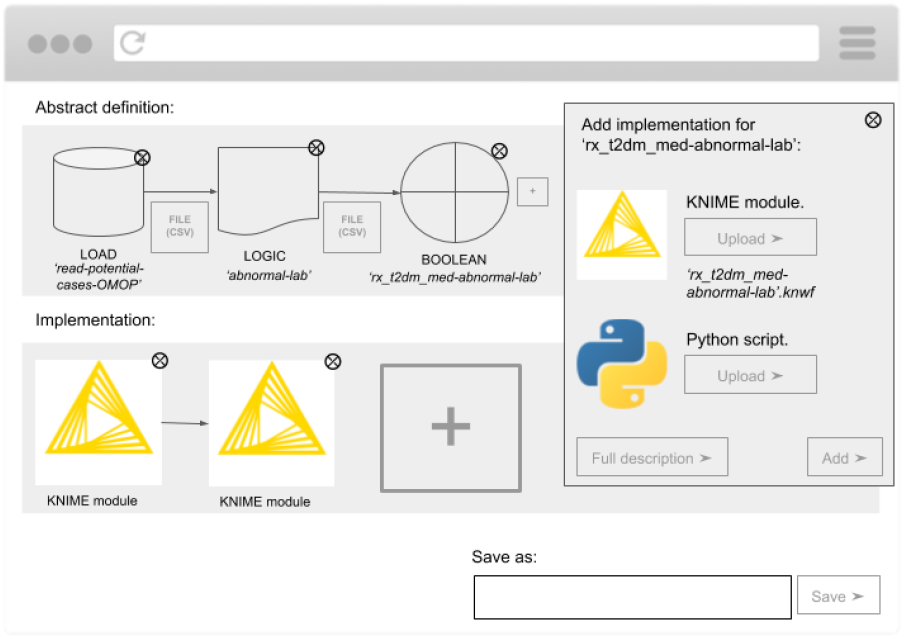
Providing an implementation unit for the third step in the abstract and functional layers, in order to generate a step in the computational layer and store this unit.

If another researcher wishes to later supply an alternative implementation unit for any of the existing units, thus introducing an additional module in the computational layer, they can do so, and this process is represented in Figure 8. Here, another author has accessed a previously authored definition, and is in the process of adding an alternative implementation for the third step in the computational layer; previously implemented as a KNIME module, the second author is now uploading a Python realisation of the same abstract boolean expression.

**Figure 8:**
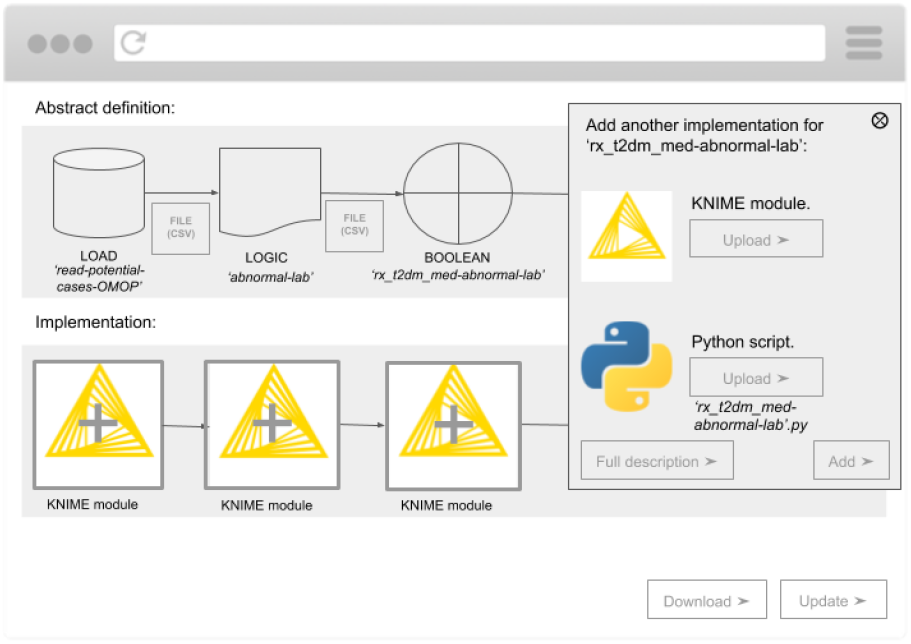
Adding additional implementation units for an existing step.

Given the potential for multiple permutations of the computational layer, and associated implementation units, when accessing the definitions authored by others, a user is able to pick the permutation they wish to use in order to generate a computable phenotype for local use. This process is represented in Figure 9, where a user is selecting, from the stored implementation units, the exact structure of the computable phenotype; they have chosen a permutation that mixes KNIME and Python implementation units.

**Figure 9:**
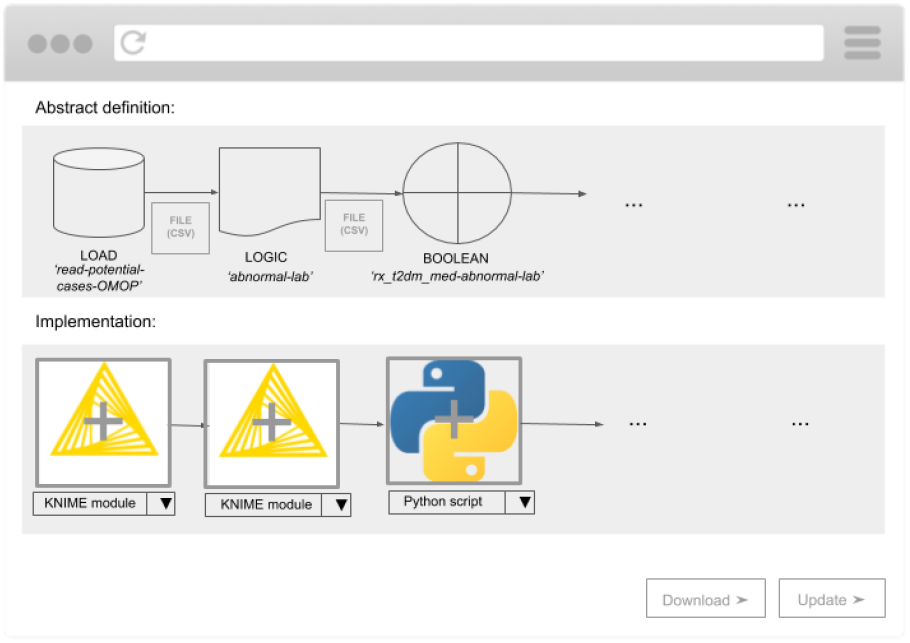
Customising a computable phenotype for local use.

Once a phenotype has been defined, and a user has customised the implementation units connected to this definition, the information elicited by the web portal is sent to the generation service in the Phenoflow architecture, which, backed by *python-cwlgen* (https://github.com/common-workflow-language/python-cwlgen), instantiates the definition as a text-based Common Workflow Language (CWL) document, and sends it back. This document is then combined with the stored implementation units, and packaged as a download for the user to execute locally as a computable phenotype, using one of CWL’s execution engines (e.g., *cwl-tool*, https://github.com/common-workflow-language/cwltool). As these engines typically integrate with container technology, we have developed several custom images to support the execution environments specified in the computational layer, including a custom KNIME Docker image.

Once it has received the CWL document back from the generator service, the web portal also sends this document to the visualisation service in the Phenoflow architecture, which, backed by *cwlviewer* (https://github.com/common-workflow-language/cwlviewer), sends back a visualisation of the abstract and functional layers expressed in the supplied workflow. This ensures that the text-based CWL instantiation of the definition is complemented by a visualisation that presents the definition in a format more commonly seen (e.g. as seen in Figure 2).

## Results

Table 1 presents the KIP portability scores for traditional code-based and logic-based phenotype definitions, and their structured counterparts. These scores were discussed and agreed upon by all of the authors, a subset of whom have extensive experience both applying and validating KIP scores. The KIP assigns a score between 0 and 2 to phenotype definitions under a number of different portability aspects, with higher scores indicating that a definition is less portable. To understand these scores better, the following sections present the impact of our model on our representative phenotype definitions for COVID-19 and T2DM, under each aspect of the KIP. Recall that we are able to directly compare the portability of the traditional and structured forms of a definition, as we have already verified that converting from one form to the other does not affect the logic of a phenotype.

**Table 1:**
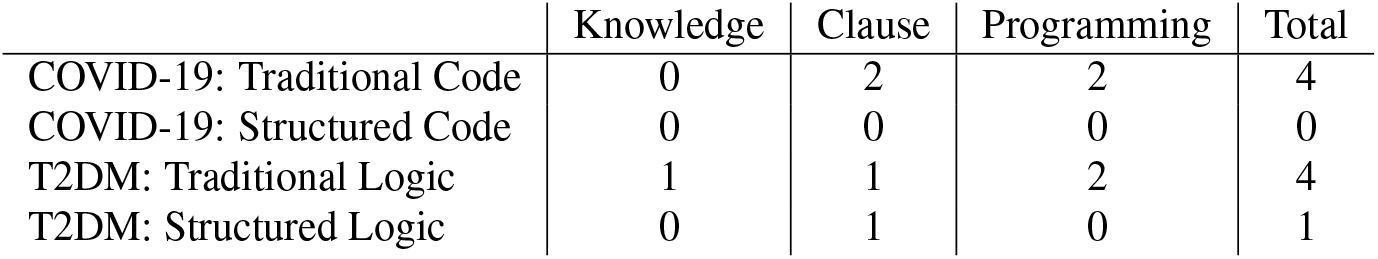
KIP scores indicating the portability of traditional code-based (COVID-19; GSTT) and logic-based (Type 2 Diabetes (T2DM); NMEDW) phenotype definitions and their structured counterparts.

## Knowledge conversion

The first aspect of the KIP scoring system relates to the clinical knowledge required to develop a computable phenotype from its definition. For example, the original COVID-19 phenotype definition is based on common vocabularies, and is thus awarded a knowledge conversion score of 0. However, in the original T2DM phenotype definition, we note the use of some more complex medical concepts (e.g., *T2DM Rx precedes T1DM Rx*, Figure 2). Supplementary information about the meaning of this terminology is provided, but not within the definition itself, and in a different form (as an additional written document). As a result, we assign a score of 1 for knowledge conversion for the original T2DM definition.

In its structured form (Figure 10), the COVID-19 definition retains its portability level for knowledge transfer, and while the T2DM phenotype (Figure 11) retains the terminology found in the original definition (e.g. *rx_t2dm_med-abnormal_lab*), the impact this has on portability is lessened in two key ways. Firstly, additional information about the meaning of the terminology is provided within the abstract layer itself, in the description field of each step, ensuring that any medical terminology is supplemented by a longer, more accessible, description (e.g. an explanation of abnormal lab values). Secondly, the classification of each step as a type of operation from a pre-defined ontology ensures that even in the presence of medical terms, basic understanding about the logic of a step can still be extracted. For example, the classification of a step containing a *case assignment* rule as a boolean expression ensures that the use of medical terminology does not obscure its logic. Based upon these factors, the KIP system assigns a value of 0 for this aspect.

**Figure 10:**
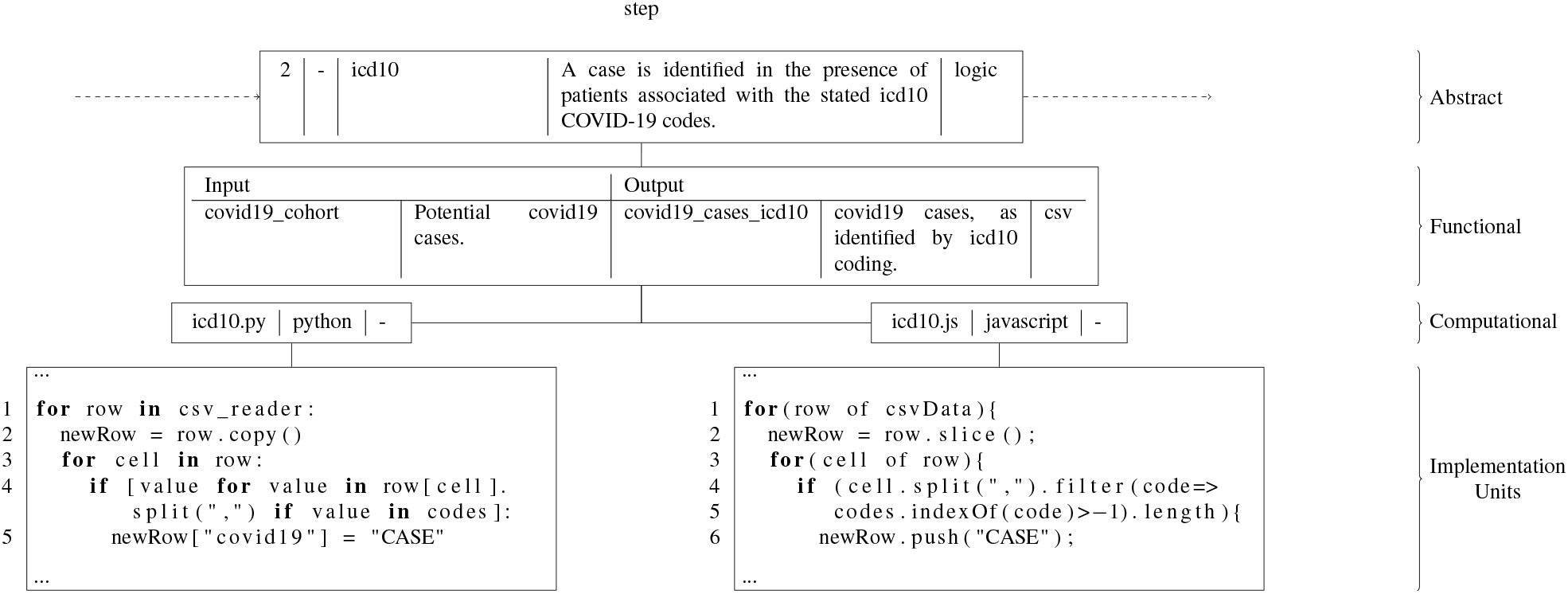
Individual step of COVID-19 structured phenotype definition and new implementation units.

**Figure 11:**
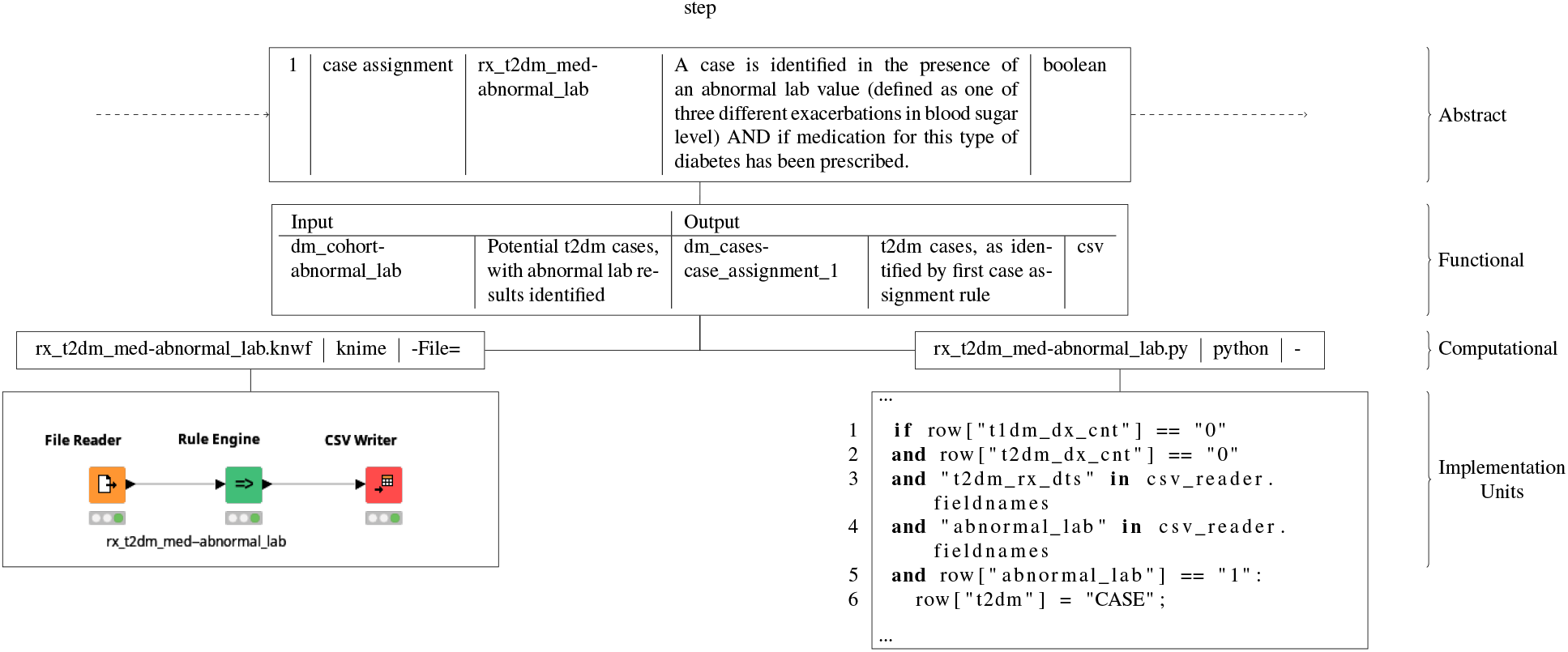
Individual step of T2DM structured phenotype definition and new implementation units.

### Clause interpretation

The second aspect of the KIP scoring system aims to identify any ambiguity in the logical clauses found in a phenotype definition, which may result in inconsistencies when realising this logic computationally. The existing T2DM phenotype definition (Figure 2) uses long conditional clauses (represented graphically), however the logic still has a clear interpretation. This leads to the attribution of a further KIP score of 1. In contrast, the COVID-19 definition, existing as a set of code lists, has a much less clear interpretation, omitting key information, such as the order in which the codes are to be applied, and how the lists are logically connected (e.g. conjunctive vs. disjunctive). This results in the awarding of a KIP score of 2 for this aspect.

The translation of the code-based COVID-19 definition to the structured form enables much of this key information to be expressed explicitly. For example, as can be seen from Figure 12 where a visualisation of the re-authored COVID-19 definition is shown, the order in which each set of codes is to be applied is now clear, and their incremental application confirms a disjunctive connection. For this reason, a new portability score of 0 is assigned. The impact of the structured form on the T2DM definition is less marked. An additional visualisation containing the abstract layer of the re-authored T2DM definition is shown in Figure 13, where the second box shows a grouping of case assignment rules as a set of nested steps, referenced by the parent step shown in orange in the first. Each of the steps in this group, which are evaluated in sequence, contains an individual boolean expression, such as the one defined in Figure 11. This aims to increase the clarity of the interpretation further, by breaking down the long clauses seen in the original abstract layer (Figure 2). Moreover, the use of a group (nested steps) itself, furthers this clarity by allowing for the overall role of these steps to be more easily identified within the abstract layer. However, while there is a simpler overall structure, at the same time the longer descriptions within each step introduce different complexity. For these reasons, the same KIP score of 1 is assigned to this aspect under the structured definition.

**Figure 12:**
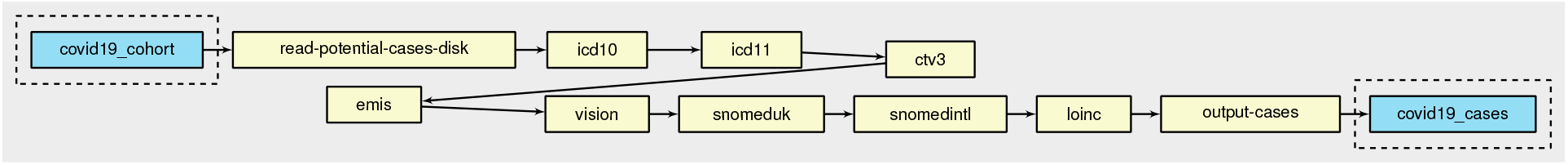
Visualisation of COVID-19 structured phenotype definition

**Figure 13:**
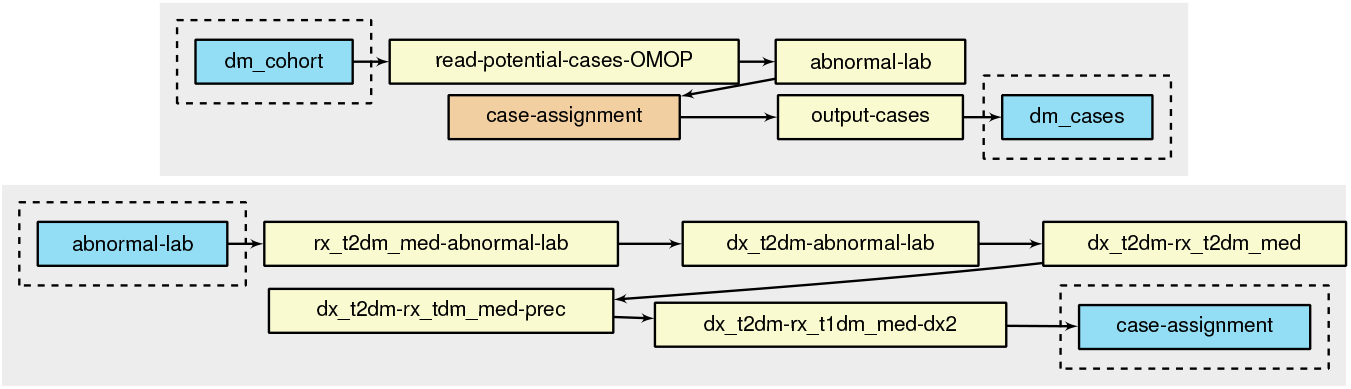
Visualisation of T2DM structured phenotype definition.

### Programming

The final aspect of the KIP scoring system relates to the programmatic complexity of implementation. The structure of the original COVID-19 definition suggests a low level of programming expertise required to produce a computable form (e.g. the requirement to produce a Python script to identify the stated codes within a dataset), while the T2DM definition suggests a moderate level of programming expertise (e.g. the requirement for a data pipeline to be produced to realise the stated logical conditions). However, the fact that little instruction can be extracted from each definition on how to develop a computable form in practice increases the complexity of implementation, and results in a disconnect between the two, which reduces the intelligibility of the implementation. This is particularly marked in T2DM, where the case assignment logic seen in Figure 2, while defined as separate operations in the definition, is obscured in a single node in the computable form (*assign case status*, Figure 3), making the correspondence between the two unclear. This reduced intelligibility makes it harder to reuse, or modify, the provided implementation in a new use case. As a result of this complexity, and the implications, a KIP score of 2 is awarded for both original definitions accordingly.

In contrast, the requirement for a distinct (set of) implementation unit(s) for each step in the abstract layer, introduced by the computational layer of a structured definition, each of which responds to the inputs, and produces the outputs, specified in the functional layer, provides a clear template for development. This lessens the implementation burden, in the case of both the COVID-19 and T2DM definitions, by either structuring new development, or allowing existing implementation units, which may have been developed locally, to be reused in order to produce the computable form of a definition.

In addition, a computable phenotype produced on the basis of a structured definition is inherently more intelligible, as the implementation holds a greater correspondence with the abstract layers. For example, in the case of T2DM, because each step in the abstract layers must be connected to an individual implementation unit, the case assignment logic is no longer obscured (as seen in Figure 11), as it was in the original computable form. As a result of this increased intelligibility, these computable phenotypes are more transparent, and thus reusable and more easy to modify, lessening the implementation burden on future developers. Moreover, assuming multiple implementation units exist for the same abstract step (previously written by other authors), which can be easily swapped in and out owing to the modularity brought by the computational layer, a user is more likely to find a unit written in a technology they are comfortable with, and can thus edit, again reducing the implementation burden. For example, our structured COVID-19 and T2DM definitions reference a mix of Python, Javascript and KNIME implementation units.

The ability to modify existing computable phenotypes structured according to our model is only increased by their delivery as executable CWL documents by the Phenoflow architecture. As CWL documents, modifications to these phenotypes can be rapidly tested against execution engines that leverage container technology to avoid having to manually install execution environments. All of these factors result in the attribution of a score of 0 for the KIP programming aspect, for both the COVID-19 and T2DM structured definitions.

## Discussion and Conclusion

In this paper, we introduce a workflow-based, multi-layer model for the definition of a phenotype, and an associated microservice architecture, *Phenoflow*, which is used to define phenotypes under this model, and export them as workflows, which can later be executed against a dataset along with associated implementation units.

Overall, we note a number of improvements to portability when a phenotype definition is structured using our representation model, under the KIP scoring system. For code-based definitions, benefits are best seen in terms of a clarity of structure, brought by the requirements of our model, while fewer improvements in portability are seen in terms of terminology. For logic-based definitions, benefits are best seen in terms of clarity of terminology, brought by supplementary information in the abstract layer, while fewer improvements are seen in terms of clause interpretation, where the fact that long clauses are (necessarily) replaced with individual steps, introduces different complexity with equivalent effect. For both code and logic definitions, significant portability improvements can be seen in terms of programmatic complexity of implementation, where a structured definition both closely guides the implementation via the additional (functional and computational) layer information, and promotes the development of intelligible computable forms that can later be reused and modified by other authors. Portability is improved even further by the presence of the Phenoflow architecture, which facilitates the collation of implementation units, and facilitates the generation of computable phenotypes from structured definitions. Ultimately, the improved phenotype portability brought about by our approach not only helps researchers reuse existing definitions in new studies, but also assists in determining the reproducibility of the methods found in published studies.

While the issue of translating an abstract phenotype definition into a computable form is well recognised within the research community, this work offers several key advancements to complement previously developed methods.

The electronic Medical Records and Genomics (eMERGE) Network has a significant record of representing phenotypes for dissemination and publication. This process was originally done by each institution within the eMERGE Network taking a narrative description of the phenotype pseudocode and an accompanying data flow diagram, and translating this into executable code that would run against their local data warehouse. This approach has now progressed to the use of pipeline-based executable representations, such as those using the KNIME analytics platform, which allows the definition of the computable form in a graphical manner. In addition, the eMERGE Network has adopted a common data model – the OMOP CDM – to facilitate the representation and dissemination of phenotype algorithms^15^. This has allowed the graphical authoring of phenotypes using the ATLAS authoring tool. This provides a human-readable representation of the logic, with the benefit of being stored in a format that may be automatically converted to an executable format across multiple database systems at different organisations. While this approach addresses the issues associated with translating an abstract definition into a compatible form and has facilitated the rapid sharing and execution of phenotypes, the OMOP CDM is not globally adopted (although it has seen wide growth and adoption in recent years), the representations are therefore not fully portable to other CDMs or local data models^16^. In contrast, phenotypes developed under our model are not tightly coupled to a single CDM; OMOP CDM is just one data source that can be referenced in a definition (as are standards such as i2b2 and Fast Healthcare Interoperability Resources (FHIR)).

The PhEMA project has also attempted to address the issue of translating an abstract definition to a computable form by proposing the use of a graphical authoring environment that can be used to generate a higher-level, standardised representation of the phenotype logic^17^. Initial work utilised the Quality Data Model (QDM), with more recent development adopting the Clinical Quality Language (CQL). A key aspect of PhEMA’s approach is the use of *translators* to take the higher-level representation (QDM or CQL), and convert it into an executable format that may run against a particular CDM. For example, the approach of converting QDM into an executable KNIME pipeline allowed that KNIME representation to still be customised for local execution^18,19^. However, while this also aims to solve the issue of developing a computable phenotype based on an abstract representation, the translators available are often specific to an implementation format, such as KNIME. In contrast, phenotypes developed under our model are not coupled to a single implementation format.

Future work will explore the impact that Phenoflow has on the portability of additional types of phenotype definitions, including probabilistic definitions, the development of which is likely to leverage data processing tools such as the Flexible Data-Driven Pipeline *(FIDDLE)* framework^20^. In addition, future work will investigate how the multidimension annotations of the structured definition model can be leveraged in order to introduce new search and discovery capabilities into phenotype repositories. For example, the ability to use a wider range of search criteria, or to understand when existing definitions intersect with those currently in a repository, to assist in finding partial phenotype matches for a user’s requirements, which can then be adapted to suit their needs. Future work will also focus on developing libraries of both abstract steps, and implementation units, to be made available to researchers wanting to customise an existing computable phenotype within Phenoflow for their research tasks. Such efforts are already underway in the UK, under the umbrella of the Health Data Research UK (HDR UK, https://www.hdruk.ac.uk) network which is developing a National Human Phenome portal, of which Phenoflow is a part. Moreover, a broader range of implementation languages will be supported, by developing implementation unit plugins, such as those that perform file type conversion, e.g., from a CSV file to a lightweight SQL table, so that an SQL script can be executed against the data within an individual step. Finally, we will investigate how our experiences of developing our structured definition model and the Phenoflow architecture can be extrapolated to a set of heuristics to be followed when designing and sharing novel phenotype definitions.

## Data Availability

This manuscript references, and is supported by, both private data from the Northwestern Medicine Enterprise Data Warehouse (NMEDW), data availabe on request from Guy's and St Thomas' Hospitals NHS Trust, and public data from PheKB.

https://www.phekb.org/phenotype/type-2-diabetes-mellitus

